# Altered cerebellar oscillations in Parkinson’s disease patients during cognitive and motor tasks

**DOI:** 10.1101/2021.08.21.21262412

**Authors:** Taylor J. Bosch, Christopher Groth, Tiffany A. Eldridge, Etienne Z. Gnimpieba, Lee A. Baugh, Arun Singh

## Abstract

Structural and functional abnormalities in the cerebellar region have been shown in patients with Parkinson’s disease (PD). Since the cerebellar region has been associated with cognitive and lower-limb motor functions, it is imperative to study cerebellar oscillations in PD. Here, we evaluated cerebellar electroencephalography (EEG) during cognitive processing and lower-limb motor performances in PD. Cortical and cerebellar EEG were collected from 74 PD patients and 37 healthy control subjects during a 7-second interval timing task, 26 PD patients and 13 controls during a lower-limb pedaling task, and 23 PD patients during eyes-open/closed resting conditions. Analyses were focused on the mid-cerebellar Cbz electrode and further compared to the mid-occipital Oz and mid-frontal Cz electrodes. Increased alpha-band power was observed during the eyes-closed resting-state condition over Oz, but no change in alpha power was observed over Cbz. PD patients showed higher dispersion when performing the 7-second interval timing cognitive task and executed the pedaling motor task with reduced speed compared to controls. PD patients exhibited attenuated cue-triggered theta-band power over Cbz during both the interval timing and pedaling motor tasks. Connectivity measures between Cbz and Cz showed theta-band differences, but only during the pedaling motor task. Cbz oscillatory activity also differed from Oz across multiple frequency bands in both groups during both tasks. Our cerebellar EEG data along with previous magnetoencephalography and animal model studies clearly show the alteration in the cerebellar oscillations during cognitive and motor processing in PD.

## Introduction

The cerebellum has been implicated in cognitive control, besides playing a pivotal role in the control and coordination of motor functions. Growing evidence has suggested structural and functional abnormalities in the cerebellum in patients with Parkinson’s disease (PD) which may contribute markedly to the cognitive and motor symptoms of PD (Pedroso *et al*., 2013; Wu and Hallett, 2013). Recent studies have shown that the cerebellar vermis and lobule V of the cerebellar anterior lobe participate actively during a dual cognitive and motor task (Wu *et al*., 2013; Gao *et al*., 2017) and suggests that the modulations in cerebellar nuclei can affect the integrated cognitive and motor networks in PD. Both the cerebellum and basal ganglia receive input from and send output to the cerebral cortex area, forming an integrated functional network whose reciprocal anatomical connections have been confirmed using retrograde transneuronal viral transport in monkeys (Bostan *et al*., 2010). These connections provide a method for abnormal signals from the basal ganglia to alter cerebellar function in PD (Bostan and Strick, 2018). Altogether, the basal ganglia, the cerebellum, and the cerebral cortex form an integrated network that can control cognitive and motor processing. However, the cerebellum is one of the least characterized and investigated brain structures in this network in PD.

Deep brain stimulation (DBS) of the basal ganglia nuclei has shown therapeutic effects in PD patients, however, the effects are mostly restricted to improved upper-limb motor performance (Pollak *et al*., 2002; Benabid, 2003; Sharma *et al*., 2020). In the advanced stage of parkinsonism, patients exhibit more debilitating lower-limb motor symptoms such as gait and balance abnormalities as well as cognitive impairment. These advanced symptoms in PD cannot be treated effectively with levodopa or high-frequency DBS approaches (Hausdorff *et al*., 2003; St George *et al*., 2010; Espay *et al*., 2012; Rocchi *et al*., 2012). Recent studies have also demonstrated the relationship between cognitive deficits and gait or lower-limb motor impairment in PD patients (Kelly *et al*., 2012). Therefore, it is critical to seek another therapeutic target that can be used to improve both cognitive and lower-limb motor functions in PD.

Neuroimaging and animal studies have shown contributions of cerebellar areas in cognitive and motor processing in normal and disease models (Liu *et al*., 2013; Wu and Hallett, 2013; Festini *et al*., 2015; Gao *et al*., 2017). However, cerebellar oscillatory activity or cerebellar electroencephalography (EEG) in humans has only been discussed in some recently published articles where they proposed that cerebellar oscillations can be a physiological signature and therapeutic target for disease models (Andersen *et al*., 2020; Pan *et al*., 2020).

Cerebellar EEG in humans can be problematic due to its limited spatial resolution for deep sources. However, some recently published articles have discussed the possibility of detecting cerebellar EEG in detail and proposed appropriate methods with a practical guideline to optimize the collection of cerebellar oscillations with EEG electrodes (Todd *et al*., 2018; Andersen *et al*., 2020; Samuelsson *et al*., 2020). In the current study, we followed all of these practical guidelines to collect signals with high signal-to-noise ratio from the mid-cerebellar vermis area by placing an EEG electrode over the posterior fossa that corresponds to medial aspects of lobules VII, VIII, and IX (Todd *et al*., 2018). Furthermore, we compared mid-cerebellar oscillations during an interval timing cognitive task and a lower-limb pedaling motor task between PD patients and healthy control subjects. Our experimental approach showed altered mid-cerebellar oscillations in PD, suggesting a pathophysiological phenomenon and potential therapeutic target to improve cognitive and motor function.

## Methods

### Participants

For all experiments, a total of 79 PD patients and 37 healthy control subjects were recruited. These patients were examined by a movement disorders physician to verify that they met the diagnostic criteria recommended by the United Kingdom PD Society Brain Bank. All PD patients and healthy control subjects provided written informed consent in accordance with the Declaration of Helsinki and the Ethics Committee on Human Research. All research protocols were approved by the University of Iowa and the University of South Dakota Human Subjects Review Boards. We performed the Montreal Cognitive Assessment (MOCA; scale from 0 to 30; lower scores are worse) to measure cognition (Nasreddine *et al*., 2005), and the freezing of gait questionnaire (FOGQ; scale from 0 to 24; higher scores are worse) to evaluate gait severity (Giladi *et al*., 2000) in PD patients.

Out of all participants, 23 PD patients for resting-state data, 74 PD patients and 37 healthy controls for the interval timing task data, and 26 PD patients and 13 healthy controls for the lower-limb pedaling task data were included (Table 1). All participants who performed the lower-limb motor task also participated in the interval timing task. We also recruited 5 PD patients to collect cerebellar EEG and electromyography (EMG) signals from the nearby muscles during resting and body actions. In our previous reports, all clinical details of the patients and control groups who participated in the cognitive task (Singh *et al*., 2021) and lower-limb pedaling motor task (Singh *et al*., 2020) are described and have been summarized in Table 1. All participants performed the interval timing cognitive and lower-limb pedaling motor tasks with levodopa medication. Our previous work has shown no effect of levodopa on the interval timing task and midfrontal low-frequency oscillations (Singh *et al*., 2021). Also, PD patients can exhibit freezing of gait which may cause fall risk during the off medication state. Therefore, to match with real-word motor function, PD patients performed the lower-limb pedaling motor task with levodopa medication (Singh *et al*., 2020).

**Table 1.** Demographic details of participants.

| **Interval timing cognitive task** |  |  |
| --- | --- | --- |
|  | **37 Controls** | **74 PD patients** |
| Gender, M/F | 21/16 | 53/21 |
| Age, years | 71.9 (1.2) | 68.6 (0.9) |
| Disease Duration, years | - | 5.1 (0.5) |
| LEDD, mg | - | 849 (53) |
| MOCA (0-30) | 26.7 (0.3) | 24.2 (0.4)* |
| mUPDRS (0-56) | - | 13.3 (0.8) |
| **Lower-limb pedaling motor task** |  |  |
|  | **13 Controls** | **26 PD patients** |
| Gender, M/F | 8/5 | 17/9 |
| Age, years | 69 (2.3) | 67.3 (1.8) |
| Disease Duration, years | - | 6.2 (0.7) |
| LEDD, mg | - | 920.8 (94.7) |
| MOCA (0-30) | 26.6 (0.5) | 23.3 (0.8)* |
| mUPDRS (0-56) | - | 14.8 (1.4) |
| FOG questionnaire (0-24) | - | 9.2 (1.2) |
| **Resting eyes-open and closed** |  |  |
|  | - | **23 PD patients** |
| Gender, M/F | - | 17/6 |
| Age, years | - | 66.8 (1.9) |
| mUPDRS (0-56) | - | 16.2 (1.3) |
| **EEG and EMG: Rest and Action** |  |  |
|  | - | **5 PD patients** |
| Gender, M/F | - | 3/2 |
| Age, years | - | 67.4 (1) |
| mUPDRS (0-56) | - | 39.8 (8.2) |

Values are expressed as mean (standard error of mean). Independent t-test was performed for comparison between controls vs PD patients. \* $p < 0.01$ . Abbreviations: Male (M); Female (F); Levodopa equivalent daily dose (LEDD); Montreal Cognitive Assessment (MOCA); motor Unified Parkinson's Disease Rating Scale (mUPDRS); Freezing of gait (FOG).

### Resting-state EEG and EMG signals analysis

Resting-state EEG data were collected during eyes-open and eyes-closed conditions to compare mid-occipital (Oz) and mid-cerebellar (Cbz) signals in PD patients (n=23). Resting-state data were epoched into consecutive epochs (5 seconds). Spectral analysis was implemented on epoched and preprocessed (see preprocessing steps below) data using the “pwelch” function in MATLAB. We selected a 1-second time window, and the number of overlapping samples was set to 50% of the window length. Since eyes-closed EEG data are associated with alpha-band (8-12 Hz) peaks in the occipital region (van Dijk *et al*., 2008; Wan *et al*., 2019) rather than the cerebellar region, we exported the amplitude of mid-occipital and mid-cerebellar alpha oscillations to study volume conduction. In addition, we performed phase analysis using wavelet method and envelope correlation analysis comparing Cbz and Oz during the eyes-open and eyes-closed conditions in the alpha-band to compare phase angle differences and correlation coefficients between both signals.

Furthermore, we recorded 180 seconds of cerebellar EEG signals from two electrodes and EMG signals from nearby muscles to compare signals in PD patients (n=5) during resting-state and action conditions. EMG electrodes were placed above the semispinalis capitis muscle. Participants performed alternating upper and lower limb flexion-extension movements for 2-3 min without any cue during the action task. We constructed a bipolar cerebellar EEG signal, fitted an envelope over the rectified high-pass (1 Hz) filtered EMG signal, and compared the two in the time-frequency domain using the “pspectrum” function in MATLAB. Further, both signals were compared in different frequency bands (5 Hz, 10 Hz, 15 Hz, and 20 Hz) via phase analysis (wavelet method), correlation coefficient (“corrcoef”), and cross-correlation (“xcorr”) methods.

### Interval timing cognitive and lower-limb pedaling motor tasks

For the interval timing task used to assess differences in cerebellar oscillations during a cognitive task, individuals were required to estimate the duration of either a 3- or 7- second interval. Briefly, individuals were instructed before the beginning of interval estimation whether the upcoming interval was short or long. Afterwards, a visual cue (solid box) on a computer screen was displayed for either 8-10 seconds for the short interval conditions or 18-20 seconds for the long interval conditions. Individuals were then required to indicate once they believed the target duration had been achieved by pressing the space bar on a computer. Here, we analyzed only 7-s interval timing task-related signals since no electrophysiological differences in frontal theta-band activity and less variation in response time were observed during the 3-s interval timing task in PD patients compared to controls (Singh *et al*., 2021). Also, in prior work, distinctions between theta-band activity were more reliable for the longer 7-second intervals than for 3-second intervals (Parker *et al*., 2015; Kim *et al*., 2017; Singh *et al*., 2021). There were 40 trials of the 7-second interval condition. To analyze cognitive behavior during the interval timing task, we computed the coefficient of variation of the response time (RTCV) (Singh *et al*., 2021).

For the lower-limb pedaling task used to assess differences in cerebellar oscillations during a motor task, a protocol previously developed to study lower-limb movement control specifically in PD patients was used (Singh *et al*., 2020). Briefly, individuals were seated and instructed to complete one full rotation of the pedaling device after being presented with a visual cue (green circle) 1-2 seconds after receiving a warning cue. This was completed between two separate blocks of either 30 or 50 trials per block. Similar to our prior report, we computed the linear speed for pedaling trials from the accelerometer signals (Singh *et al*., 2020).

### Cerebellar and cortical EEG signals and analysis

In the current study, we used a customized 64-channel EEG cap with embedded active electrodes (actiCAP, EasyCap, Inc) in accordance with the extended version of the international 10/20 system. EEG signals were collected using a high pass filter of 0.1 Hz and a sampling rate of 500 Hz. During data collection, the Pz electrode was used as the reference electrode and the FPz electrode was used as the ground electrode. This EEG cap was customized to include electrodes covering the posterior fossa corresponding to the medial aspects of cerebellar lobules VII, VIII, and IX. The I1 and I2 electrodes corresponding to cerebellar (Cb1 and Cb2) coverage replaced PO3 and PO4 in the EEG cap. Another channel was added corresponding to a central cerebellar location (Cbz). Cerebellar electrodes were placed 1 cm below the occipital electrode (Edagawa and Kawasaki, 2017). The channels Fp1, Fp2, FT9, FT10, TP9, and TP10 were removed from subsequent analyses since they often correspond to eyeblink and muscle artifact.

Following data collection, data were imported into EEGLAB for preprocessing (Delorme and Makeig, 2004). For each task, bad channels were identified, removed, and interpolated. Following interpolation, data were epoched based on the task. For the interval timing task, EEG data were epoched starting two seconds before stimulus onset and ending twenty seconds after stimulus onset for the 7-second interval condition. For the lower limb pedaling task, data were epoched starting one second prior to stimulus onset and ending three seconds after stimulus onset. For both tasks, bad epochs were identified and rejected using a combination of the FASTER (Nolan *et al*., 2010) and ADJUST (Mognon *et al*., 2011) algorithms, and the “pop_rejchan” function in EEGLAB. These datasets then underwent independent component analysis and eye blink artifact was identified and removed by a trained experimenter.

Following preprocessing, time-frequency analyses were performed for all tasks using complex Morlet wavelets, as explained previously (Singh *et al*., 2018; Singh *et al*., 2019; Singh *et al*., 2021). Briefly, wavelets increased from 1-50 Hz in logarithmically spaced steps defining the cycles of each frequency band increasing from 3-10 cycles between 1-50 Hz and taking the inverse fast Fourier transform. Epochs were defined differently based on the task. For the interval timing task, epochs were defined starting 500 milliseconds prior to stimulus onset continuing to 1000 milliseconds after stimulus onset. For the lower limb pedaling task, epochs were defined starting 500 milliseconds prior to stimulus onset continuing to 2000 milliseconds after stimulus onset. Power was normalized by converting to a decibel (dB) scale which allows direct comparisons across frequency bands. As in our previous studies, the baseline for each frequency was calculated by averaging power from −300 – −200 milliseconds prior to stimulus onset (Singh *et al*., 2018; Singh *et al*., 2019; Singh *et al*., 2020).

Time-frequency region of interest (tf-ROI) analyses during cognitive and motor tasks were performed primarily at the electrode of interest, Cbz over the mid-cerebellar (vermis) region. Furthermore, we analyzed a nearby mid-occipital electrode, Oz, and compared between both electrodes to rule out contamination due to volume conduction. In accordance with our previous cognitive and pedaling task-related EEG reports, tf-ROI analyses were constrained to a pre-defined theta-band (4-7 Hz) and cue-triggered 0-500 milliseconds time window (Singh *et al*., 2018; Singh *et al*., 2019; Singh *et al*., 2020; Singh *et al*., 2021). In addition to this well-motivated tf-ROI, we used a cluster size of 500 pixels and a t-test as in our past work (Singh *et al*., 2020; Singh *et al*., 2021).

Furthermore, we selected a pre-defined delta-band (1-4 Hz) and our time-window of interest was 0-500 milliseconds from the cue for both cognitive and pedaling motor tasks. We also included the beta-band (13-30 Hz) in the analysis but the time-window of interest for the interval timing task was 0-500 milliseconds from the response, and 500-2000 milliseconds from the cue for the pedaling motor task. We compared power values in those frequency bands to determine the difference between mid-cerebellar Cbz and mid-occipital Oz electrodes.

We also performed task-related connectivity analyses to observe the oscillatory relationship between the mid-frontal and mid-cerebellar regions. Since our previous reports have shown abnormal midfrontal activity in the theta-band at the time of the cue during both interval timing cognitive and lower-limb pedaling motor tasks in PD patients (Singh *et al*., 2020; Singh *et al*., 2021), this analysis may shed light on the role of the cortico-cerebellar network in cognitive processing and lower-extremity movements in PD patients. We applied phase and power-based connectivity analysis methods to measure spectral coherence and power correlation between signals collected from Cbz and Cz electrodes during the interval timing cognitive and lower-limb pedaling motor tasks (Cohen, 2014).

### Statistical analysis

For the initial resting-state EEG analysis comparing mid-occipital electrode Oz to mid-cerebellar electrode Cbz, a 2×2 repeated measures analysis of variance (rmANOVA) was performed with condition (eyes-open and eyes-closed) and electrode (Oz and Cbz) as primary factors for alpha oscillations. Independent t-tests were performed to assess the difference between PD patients and healthy control subjects for the interval timing and lower-limb pedaling tasks. We also used Pearson correlation analyses to evaluate the relationship between theta and beta power values during the interval timing task and cognitive measurements (MOCA scores); and between theta and beta power values during the lower-limb pedaling motor task and gait severity measurements (FOG scores) of PD patients. Moreover, we performed an rmANOVA including electrodes (mid-cerebellar Cbz and mid-occipital Oz) as a within-subjects variable and group as a between-subjects variable (PD and controls). An alpha level of <0.05 was adjusted for multiple comparisons via the Bonferroni correction method, where appropriate. The horizontal lines and white circles in the violin plots represent the mean and median values, respectively.

## Results

### Feasibility of cerebellar EEG

Initially, we explored whether cerebellar EEG is a feasible method for indirectly assessing cerebellar activation in human subjects. Due to volume conduction and the proximity to occipital electrodes, it was important to distinguish between Cbz and Oz during resting-state EEG in PD patients (Fig. 1). The results of the 2×2 rmANOVA examining effects of electrode and resting-state condition in the alpha-band revealed a main effect of electrode (F_1,22_ = 6.56, p = 0.018), a main effect of resting-state condition (F_1,22_ = 8.56, p = 0.008), and an interaction between these factors (F_1,22_ = 4.85, p = 0.038). Subsequent pairwise comparisons demonstrated a significant difference between eyes-closed and eyes-open conditions in the mid-occipital Oz electrode (p = 0.012). Similar to previous studies, our mid-occipital EEG signal showed a regional peak in the alpha-band during the eyes-closed condition (mean ± sem = 4.14 ± 1.1 μV^2^; Fig. 1) compared to the eyes-open condition (1.65 ± 0.37 μV^2^; Fig. 1) (van Dijk *et al*., 2008; Wan *et al*., 2019). Further, alpha-band mid-cerebellar Cbz oscillations showed lower power (2.39 ± 0.48 μV^2^; Fig. 1) and were significantly different from mid-occipital Oz alpha oscillations (p = 0.02) during the eyes-closed condition, thus showing an absence of volume conduction from alpha oscillations generated in the occipital region. No difference between Oz and Cbz alpha-band power during the eyes-open condition was observed (p = 0.2). In addition, phase and envelop correlation analyses comparing Cbz and Oz during the eyes-open and eyes-closed conditions in the alpha-band showed greater and dispersed phase angle differences as well as lower and distributed envelop correlation coefficients during the eyes-closed condition (Fig. S1).

**Fig. 1.**
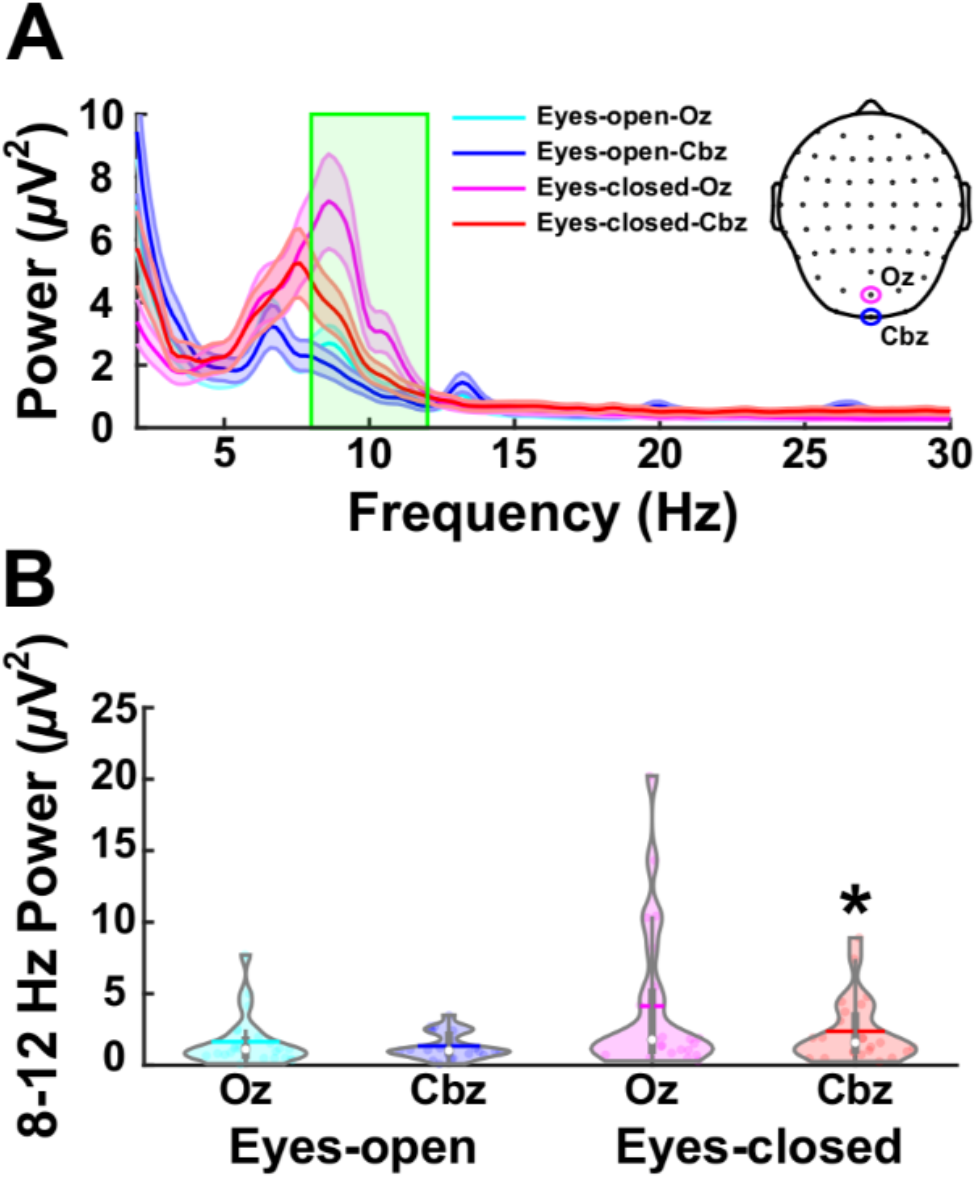
Comparison between mid-cerebellar Cbz and mid-occipital Oz alpha-band oscillations during resting-state eyes-open and eyes-closed conditions in PD participants (n = 23). A-B) Power spectral density comparing Cbz and Oz activity during resting-state EEG shows increased alpha power (8-12 Hz) during eyes closed resting-state EEG for Oz compared to Cbz. *p<0.05 vs. eyes closed Oz. The horizontal lines and white circles in the violin plots represent the mean and median values, respectively.

Moreover, signals recorded from the cerebellar EEG electrodes and nearby EMG leads during the rest and action conditions showed differences after applying 1-50 Hz band-pass filters (Fig. S2A). When enveloped signals were compared between EEG and EMG electrodes at different frequency bands (5, 10, 15, 20 Hz), phase analysis showed normally distributed phase differences (Fig. S2B), correlation coefficients were minimal between (Fig. S2C), and cross-correlation analyses showed minimal zero-lag relationships between both signals (Fig. S2D). Though activity can be seen around 5 Hz and its higher harmonics in these analyses between EEG and EMG signal, they were thoroughly preprocessed and removed in the resting-state, cognitive, and motor tasks. Altogether, both our resting-state eyes-open and eyes-closed data showed the feasibility of cerebellar EEG signals and revoked the probability of volume condition from the nearby occipital electrode. Also, cerebellar EEG and EMG signals showed that in our setup, muscle activity close to cerebellar electrodes did not contaminate the cerebellar EEG signals during the task conditions.

### Cerebellar EEG in PD patients during a cognitive task

Following the initial comparisons between Oz and Cbz oscillatory activities during resting state, we explored whether there were differences in mid-cerebellar EEG activity between PD and healthy control subjects during 7-second interval timing cognitive processing (Fig. 2A). Our recently published study on the same dataset demonstrated higher RTCV in PD patients compared to controls (RTCV: controls = 0.15 ± 0.01, PD patients = 0.24 ± 0.02; t_109_ = −3.31, p = 0.001). Behavior response outcomes clearly demonstrated cognitive deficits in our patients compared to controls. Electrophysiological activity in the mid-cerebellar region during the 7-s interval timing task showed significantly reduced cue-triggered theta-band power in PD patients compared to healthy controls (mean ± sem: controls = 1.15 ± 0.26 dB, PD patients = −0.01 ± 0.17 dB; t_109_ = 3.75, p = 0.0003; Fig. 2A-C). Interestingly, we found that cue-triggered mid-cerebellar theta activity in the interval timing task correlated with cognitive impairment in PD (r=0.28, p=0.017; Fig. 2D) and a trend was observed between theta activity and RTCV (r=−0.22, p=0.06). In addition, beta-band power in PD patients was higher compared to healthy controls (controls = −1.18 ± 0.09 dB, PD patients = −0.89 ± 0.06 dB; t_109_ = −2.52, p = 0.01; Fig. S3 A and B) and we did not find a significant correlation between beta power and MOCA scores or RTCV (Fig. S3 C and D).

**Fig. 2.**
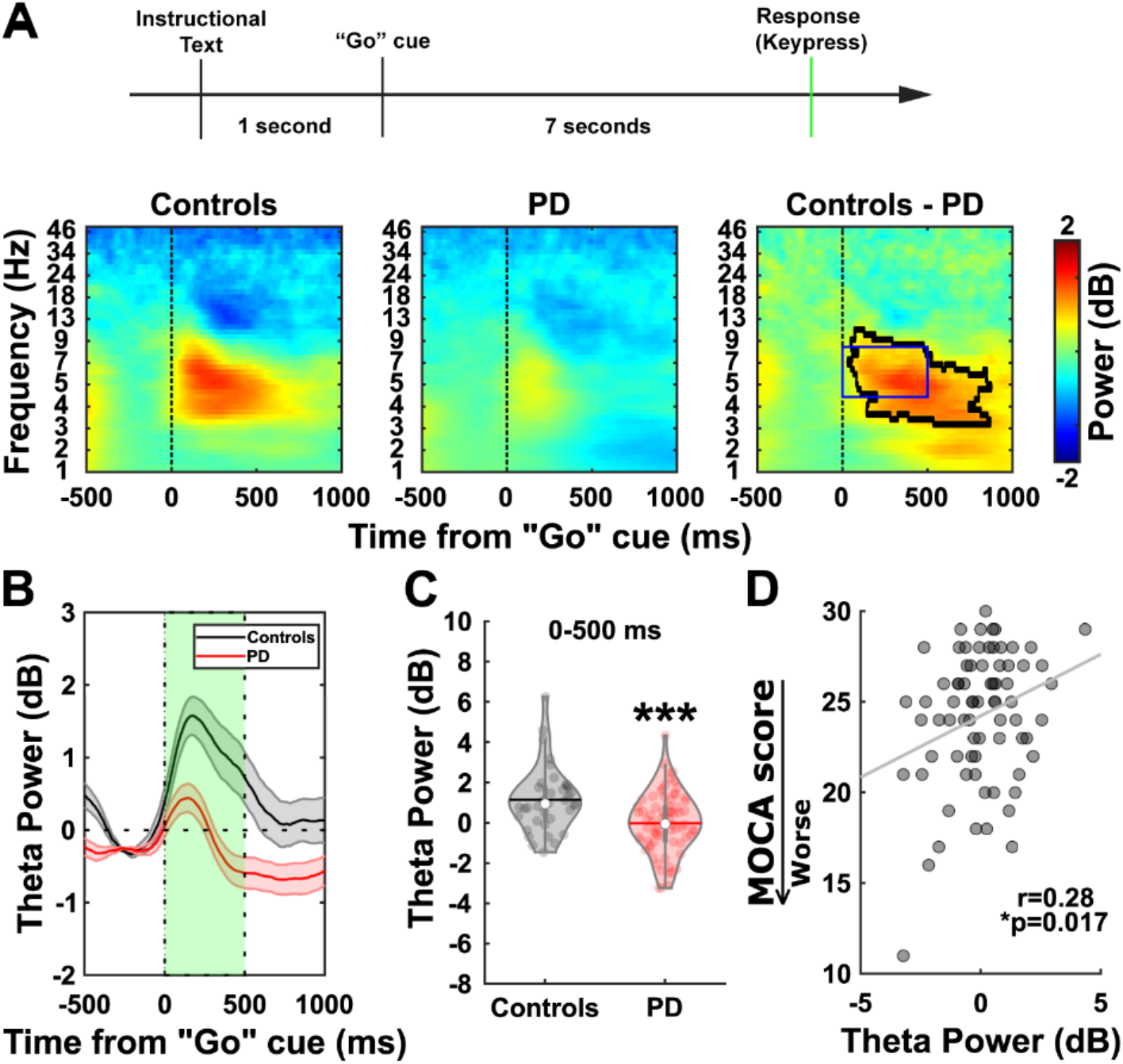
Decreased mid-cerebellar Cbz theta correlates with cognitive dysfunction in PD. A) In this task, participants were shown a “Go” cue (solid box) that was displayed for 18-20 seconds. Individuals were required to indicate when they believed the target duration (7 seconds) had passed with a keypress (top). Time-frequency analyses comparing PD and control subjects beginning at the onset of the target “Go” cue (bottom). B) Theta power (4-7 Hz) comparison between PD and control subjects beginning at the onset of the target “Go” cue. C) Theta power is significantly decreased in PD patients (tf-ROI: blue box in A and green box in B). D) Decreased theta power associates with poor cognition as indicated by the Montreal Cognitive Assessment (MOCA) scores where lower scores indicate worse cognition. ***p<0.001 vs. controls. The horizontal lines and white circles in the violin plots represent the mean and median values, respectively.

We further compared the difference between mid-cerebellar Cbz activity and mid-occipital Oz activity during the 7-second interval timing cognitive task (Fig. S4) in delta, theta, and beta frequency bands in PD patients and controls. Cue-triggered delta and theta power were significantly attenuated in the mid-cerebellar Cbz region compared to the mid-occipital Oz region in PD patients (Fig. S4 and Table S1). Furthermore, response-related beta power was significantly reduced in PD patients compared to controls during the interval timing task and a trend towards a reduction in beta power was also found in the mid-cerebellar Cbz region compared to the mid-occipital Oz region in PD patients (Fig. S4 and Table S1).

Finally, we explored the connection between mid-frontal and mid-cerebellar regions. Both spectral coherence and power correlation analyses showed no significant differences in connectivity at the cue-triggered theta-band during the interval timing task (Fig. S5).

### Cerebellar EEG in PD patients during a lower-limb pedaling motor task

Finally, we explored whether there are differences in mid-cerebellar Cbz activity between PD patients and healthy control subjects during lower-limb pedaling motor performance (Fig. 3A). Our prior study on the same dataset showed that PD patients performed the pedaling motor task with reduced speed compared to controls (speed: controls = 0.016 ± 0.002 m/s, PD patients = 0.008 ± 0.001 m/s; t_37_ = 4.2, p = 0.0002) (Singh *et al*., 2020). Mid-cerebellar EEG activity during the lower-limb pedaling motor task showed significantly reduced cue-triggered theta-band power in PD patients compared to healthy controls (mean ± sem: controls = 3.3 ± 0.67 dB, PD patients = 0.62 ± 0.27 dB; t_37_ = 4.45, p = 0.0001; Fig. 3 A-C). Similarly, our prior study demonstrated a reduction in cue-triggered mid-frontal theta power in PD patients due to participants performing the pedaling motor task after seeing the imperative stimulus, which required attention or cognitive load (Singh *et al*., 2020). Strikingly, we found that cue-triggered mid-cerebellar theta activity in the lower-limb pedaling motor task correlated with gait impairment in PD (r=−0.53, p=0.006; Fig. 3D). In addition, we compared cue-triggered beta-band power in PD patients to healthy controls but found no significant difference between groups, although the beta power was higher in the mid-cerebellar electrode in PD (mean ± sem: controls = −0.04 ± 0.2 dB, PD patients = 0.3 ± 0.2 dB; t_37_ = −1.12, p = 0.27; Fig. 3 E-F). We also noticed no association between mid-cerebellar beta-band power and gait impairment in PD (r = 0.13, p = 0.534; Fig. 3G), suggesting that changes in the beta-band during movement and the relationship between beta power and motor symptoms in PD may be limited to motor cortical, frontal, and subcortical regions (Singh *et al*., 2013; Singh, 2018; Singh *et al*., 2020).

**Fig. 3.**
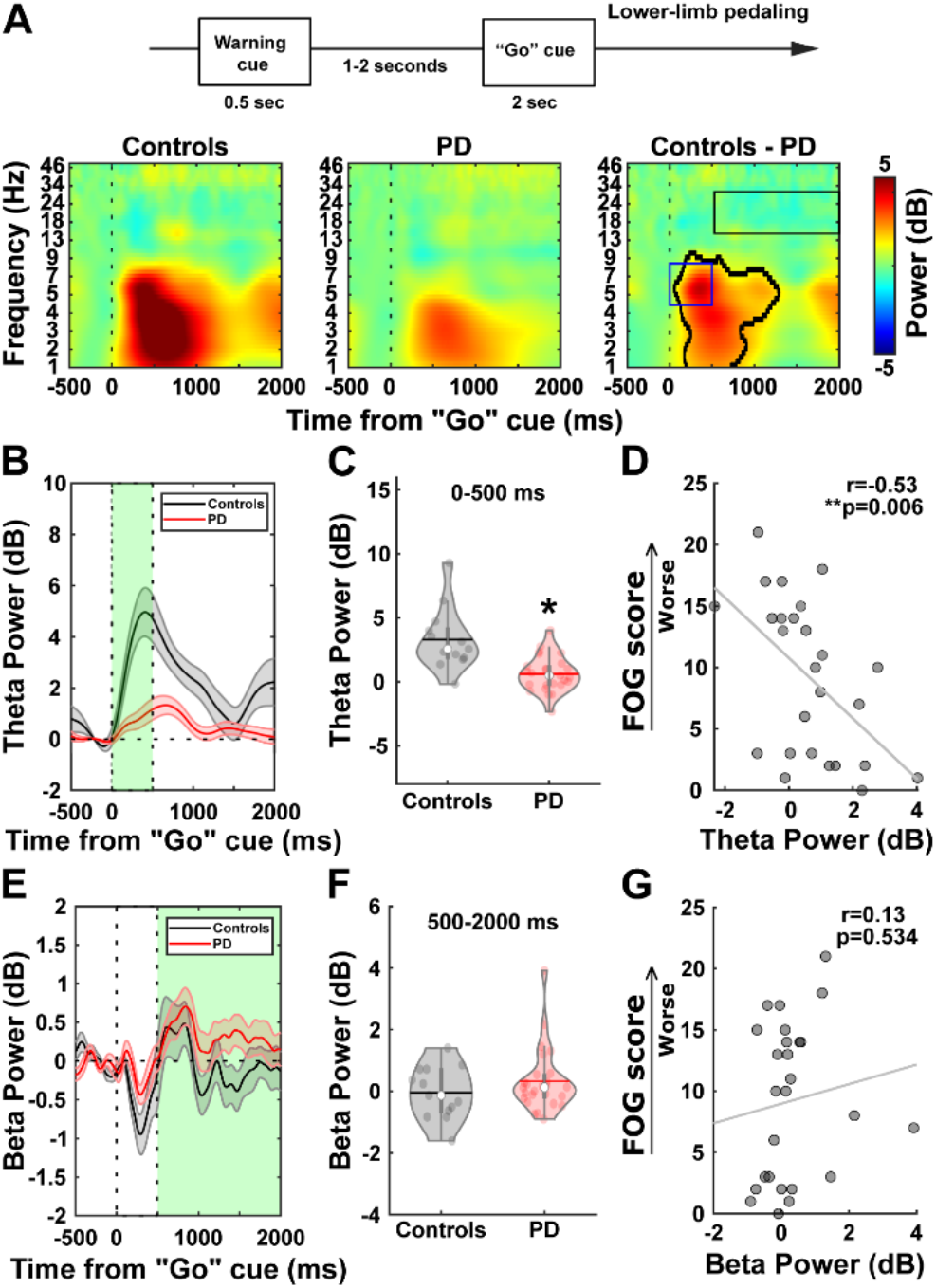
Decreased mid-cerebellar Cbz theta correlates with lower-limb motor impairment in PD. A) In this task, participants were shown a “Go” cue and were required to complete one full rotation of the pedal (top). Time-frequency analyses comparing PD and control subjects beginning at the onset of the target “Go” cue (bottom). B) Theta power (4-7 Hz) comparison between PD and control subjects beginning at the onset of the target “Go” cue. C) Theta power (4-7 Hz) is significantly decreased in PD patients (tf-ROI: blue box in A and green box in B). D) Decreased theta power associates with gait impairment as indicated by freezing of gait questionnaire (FOG) scores where higher scores indicate worse gait impairment. E) Beta power (13-30 Hz) comparison between PD and control subjects after the onset of the target “Go” cue (tf-ROI: black box in A and green box in E). F) Beta power shows no significant difference in PD patients. G) Beta power is not associated with gait impairment. *p<0.05 vs. controls. The horizontal lines and white circles in the violin plots represent the mean and median values, respectively.

Subsequently, we compared the difference between mid-cerebellar Cbz activity and mid-occipital Oz activity during the lower-limb pedaling motor task (Fig. S6) in delta, theta, and beta frequency bands in PD patients and controls. Cue-triggered delta and theta power values were significantly attenuated in both Cbz and Oz regions of PD patients compared to controls (Fig. S6 and Table S1). However, movement-related mid-cerebellar Cbz beta power was significantly elevated in PD patients compared to the mid-occipital Oz region (Fig. S6 and Table S1).

Furthermore, spectral coherence and power correlation analyses showed significant differences in connectivity at the theta-band during the lower-limb pedaling motor task (Fig. S7). Additionally, since a previous study suggested a higher expected signal from lateral cerebellar regions (Samuelsson *et al*., 2020), we further analyzed left cerebellar Cb1 and right cerebellar Cb2 signals during the interval timing cognitive and lower-limb pedaling motor tasks. Our results showed similar oscillatory changes between PD and controls in both lateral cerebellar EEG signals and mid-cerebellar EEG during both the interval timing cognitive (Fig. S8) and lower-limb pedaling motor tasks (Fig. S9). Furthermore, rmANOVAs revealed a significant main effect of electrode when comparing Cb1, Cbz, and Cb2 in the theta-band for both tasks. Table S2 and Table S3 show the mean values and rmANOVA results, respectively. These results suggest that there are differences across cerebellar electrodes in the theta-band, but differences between PD patients and controls persist across cerebellar electrodes.

## Discussion

The purpose of the following study was three-fold. First, we sought to demonstrate the viability of cerebellar EEG measurements via appropriate methods and experimental setup (Edagawa and Kawasaki, 2017; Pan *et al*., 2020; Samuelsson *et al*., 2020). Second and third, we aimed to characterize the EEG signature of cerebellar abnormalities in PD in the cognitive domain using an interval timing task (second), as well as in the motor domain using a lower-limb pedaling motor task (third). Our results demonstrate the viability of cerebellar EEG and characterize theta-band PD abnormalities in the cognitive domain and motor domain at the time of attention or cognitive load.

Since there is a well-documented alpha oscillation in the occipital region during resting-state eyes-closed conditions (van Dijk *et al*., 2008; Wan *et al*., 2019), we also confirmed the presence of a mid-occipital alpha peak in our patients during the eyes-closed condition only. However, this peak was absent over the mid-cerebellar electrode, suggesting that mid-cerebellar EEG was not affected by occipital EEG due to volume conduction. In our study, the eyes-closed related occipital alpha peak in PD patients was also unaffected by age and disease, since the alpha peak has been seen in older healthy people in resting-state eyes-closed conditions (Barry and De Blasio, 2017). Overall, our data demonstrates that the signal measured from the mid-cerebellar electrode functionally differs from the signal measured from the mid-occipital electrode.

Our interval timing task-related results provide insight into the role of cerebellar oscillations in the nature of temporal cognitive dysfunction in PD patients. First, the cerebellar network has been shown to support resonance within the low-frequency theta-band (D’Angelo *et al*., 2001; Dugue *et al*., 2009). Moreover, there is evidence in animal models that these low-frequency oscillations synchronize within cerebellar hemispheres as well as between cerebellar and cortical regions (Hartmann and Bower, 1998; O’Connor *et al*., 2002). Previous studies have suggested a relationship between cognitive processing and frontal low-frequency oscillatory activity (Cavanagh and Frank, 2014; Singh *et al*., 2018) as well as cerebellar activity (Wu *et al*., 2013; Gao *et al*., 2017). It should be noted that our prior report on the same patients has shown attenuated frontal theta activity during the interval timing task, thus suggesting the presence of altered theta oscillations in the frontal-cerebellar network (Singh *et al*., 2021). Studies on cerebellar theta oscillations indicate that our method may be engaging resonant frequencies in cerebellar networks that favor cognitive processing. Moreover, neuromodulation in the cerebellar region can increase theta activity and thus improve cognitive functions (Schutter and van Honk, 2006; Singh *et al*., 2019), possibly by inducing long-lasting changes in the interconnected cortico-cerebellar networks in the disease condition (Chen and Desmond, 2005; Koch, 2010; Brissenden *et al*., 2018).

Low-frequency theta oscillations can engage not only cortical and cerebellar regions, but also basal ganglia regions such as the subthalamic nucleus as well as other brain areas (Herz *et al*., 2016; Kelley *et al*., 2018). The substantial disynaptic projection between the subthalamic nucleus and cerebellar cortex forms an integrated functional network and provides a method for communication (Bostan *et al*., 2010). Our data suggest that in cognitively impaired PD patients, theta power is decreased, resulting in a failure to engage in temporal processing by the cortico-basal ganglia-cerebellar networks (Milardi *et al*., 2019). Our findings in PD patients and controls extend this by showing cue-triggered mid-cerebellar theta oscillations that are accompanied by cognitive engagement and coordinated with frontal cortical and basal ganglia theta rhythms. Interestingly, spectral coherence and power correlations between mid-frontal and mid-cerebellar regions in the theta-band during cognitive processing showed no differences between PD patients and healthy controls, suggesting that theta differences at frontal and cerebellar sites may be driven by similar, yet distinct underlying mechanisms.

Besides frontal low-frequency oscillations, beta oscillations have also been shown to be associated with the time of execution in cognitive tasks (Stoll *et al*., 2016) and linked to top-down behavioral control (Buschman and Miller, 2007). Intracranial and scalp EEG recordings in humans have shown changes in the beta-band power within the frontal cortex and subthalamic nucleus when responding during a cognitive processing task, which can be related to the motor component of cognitive task performance (Aron *et al*., 2007; Swann *et al*., 2011). This suggests that the beta-band could also be important for communication in the frontal-basal ganglia circuit during cognitive-motor processing. These oscillations could represent the interaction between the frontal-basal ganglia network and the cerebellar region, as shown by the beta activity at the time of response or keypress in our interval timing task data.

Besides in high-level cognitive processes, the cerebellum also plays a fundamental role in controlling and executing voluntary movement. The cerebellum’s primary motor role is to modulate frontal cortical and motor cortical regions through cerebello-thalamo-cortical connections (Strick, 1985). Our prior report has demonstrated a reduction in cue-triggered frontal theta activity during the lower-limb pedaling motor task in PD patients compared to controls (Singh *et al*., 2020). Of note, our current results in the same patients show a reduction in cue-triggered mid-cerebellar theta activity during the pedaling task in PD patients. As discussed above, the entrainment of cue-triggered theta oscillations occurs from frontal to cerebellar regions via cortico-cerebellar circuitry, specifically when one plans to execute a motor task that requires cognitive engagement. Though changes in cerebellar activity can directly affect motor control, changes in attentional and executive functions can result in slowness during lower-limb motor performance in PD. As such, attentional and other cognitive training methods have been used as an intervention to improve lower-limb movements in PD (Walton *et al*., 2018). Cerebellar recordings in animal models support our motor-task-related results by showing changes in theta activity during movements (O’Connor *et al*., 2002; Courtemanche *et al*., 2013). Indeed, entrainment of theta oscillations in the cerebellar region via theta frequency stimulation has shown improvement in visuo-motor tasks by modulating neural activity in focal and interconnected motor networks (Koch *et al*., 2020). This ability of the cerebellar region to generate theta oscillations during lower-limb movements, similar to the frontal region (Singh *et al*., 2020), suggests that these rhythms may represent a common spatiotemporal code for the fronto-cerebellar dialog. Indeed, our connectivity analyses between mid-frontal and mid-cerebellar regions in the theta band showed diminished spectral coherence and power correlations in PD patients compared to healthy controls. Our prior and current results propose that changes in theta activity across the fronto-cerebellar network may underlie deficits in lower-limb motor performance in PD patients.

Similar to our cerebellar EEG results, cerebellar MEG data in humans and cerebellar field potentials in monkeys have demonstrated modulation of beta oscillations during motor performance (Courtemanche *et al*., 2002; Herrojo Ruiz *et al*., 2017). Evidence has shown a relationship between desynchronized motor cortical beta activity and improved motor performance in PD (de Hemptinne *et al*., 2015; Singh, 2018). In addition, increased cortical and basal ganglia beta oscillations are associated with decreased movement in PD patients (Singh and Botzel, 2013; Singh *et al*., 2013; Toledo *et al*., 2014; Singh, 2018). Also, increased beta activity in the cerebellar region shows that synchronized beta activity in the cortico-basal ganglia-cerebellar network promotes tonic activity that can diminish lower-limb motor performance. In turn, this implicates abnormal beta oscillations in lower-limb motor dysfunction. However, our results showed no difference in beta activity between PD patients and healthy control subjects, suggesting that abnormal beta oscillations in lower-limb motor dysfunction may not manifest in cerebellar EEG. These results were further supported by our connectivity analyses between mid-frontal and mid-cerebellar regions in the beta band showing no differences in spectral coherence or power correlations between PD patients and healthy control subjects.

Potential limitations of the interval timing cognitive task, lower-limb pedaling motor task, and both tasks-related cortical EEG have been discussed extensively in our prior reports (Singh *et al*., 2020; Singh *et al*., 2021) since cortical and cerebellar EEG data were collected from the same participants and under similar conditions. In addition, there were several limitations to this study. EEG has low spatial resolution, so the detailed location of the cerebellum and occipital regions within the brain could not be identified. However, we confirmed that signals collected from the cerebellar electrode were different from the occipital electrode. In addition, our connectivity analyses distinguished between mid-cerebellar and mid-frontal theta oscillations in PD during our cognitive task. Future research would benefit from further examining the different contributors to cerebellar theta oscillations during cognitive processing compared to lower-limb motor processing, as our data showed abnormal cortico-cerebellar connectivity during the lower-limb motor task, but not the cognitive task. Furthermore, we are exploring intracranial cerebellar recordings in animal models and applying combined fMRI-EEG experiments to further elucidate the cerebellar basis of oscillatory abnormalities in PD during cognitive and motor processing.

Finally, given the proximity of mid-cerebellar electrodes to the cervical neck muscles, it could be argued that the differences observed in our study could be contaminated by these muscles. However, our data were visually inspected, and all practical guidelines previously outlined were followed to ensure high signal to noise ratio from our recording electrodes and all potential muscle-related artifacts were attenuated during the preprocessing steps (Todd *et al*., 2018; Andersen *et al*., 2020; Samuelsson *et al*., 2020). Furthermore, the differences observed in the theta-band, but not the beta-band, suggests that general EMG contamination did not affect our results since theta differences were examined prior to movement onset. Nonetheless, future research would benefit from simultaneously recording EMG data from nearby muscles with MEG or fMRI methods during cognitive and motor tasks to definitively rule out their influence.

## Data Availability

Data and codes will be available upon request from the corresponding author.

## Acknowledgments

We would like to thank Arturo Espinoza from the University of Iowa for his help in the data collection. TJB, TAE, and AS are supported by the Division of Basic Biomedical Sciences and Center for Brain and Behavior Research at the University of South Dakota, Vermillion, SD, USA.

## Author Contributions

Study design and concept: C.G., E.G.Z., L.A.B., A.S.; data collection: T.J.B., T.A.E.; A.S.; data analysis: T.J.B, E.G.Z., A.S.; drafting and revision of the manuscript: T.J.B., C.G., T.A.E., E.G.Z., L.A.B., A.S.

## Data Availability

All the raw data related to the interval timing task and lower-limb pedaling motor task can be found at PRED+CT (predict.cs.unm.edu). In addition, all the postprocessed data that support the findings, and matlab codes to generate figures are available upon reasonable request from corresponding author.

## Supplementary Data

Additional supplementary information can be found in the online version of this article at the publisher’s website.

## Supplementary Information

**Fig. S1.**
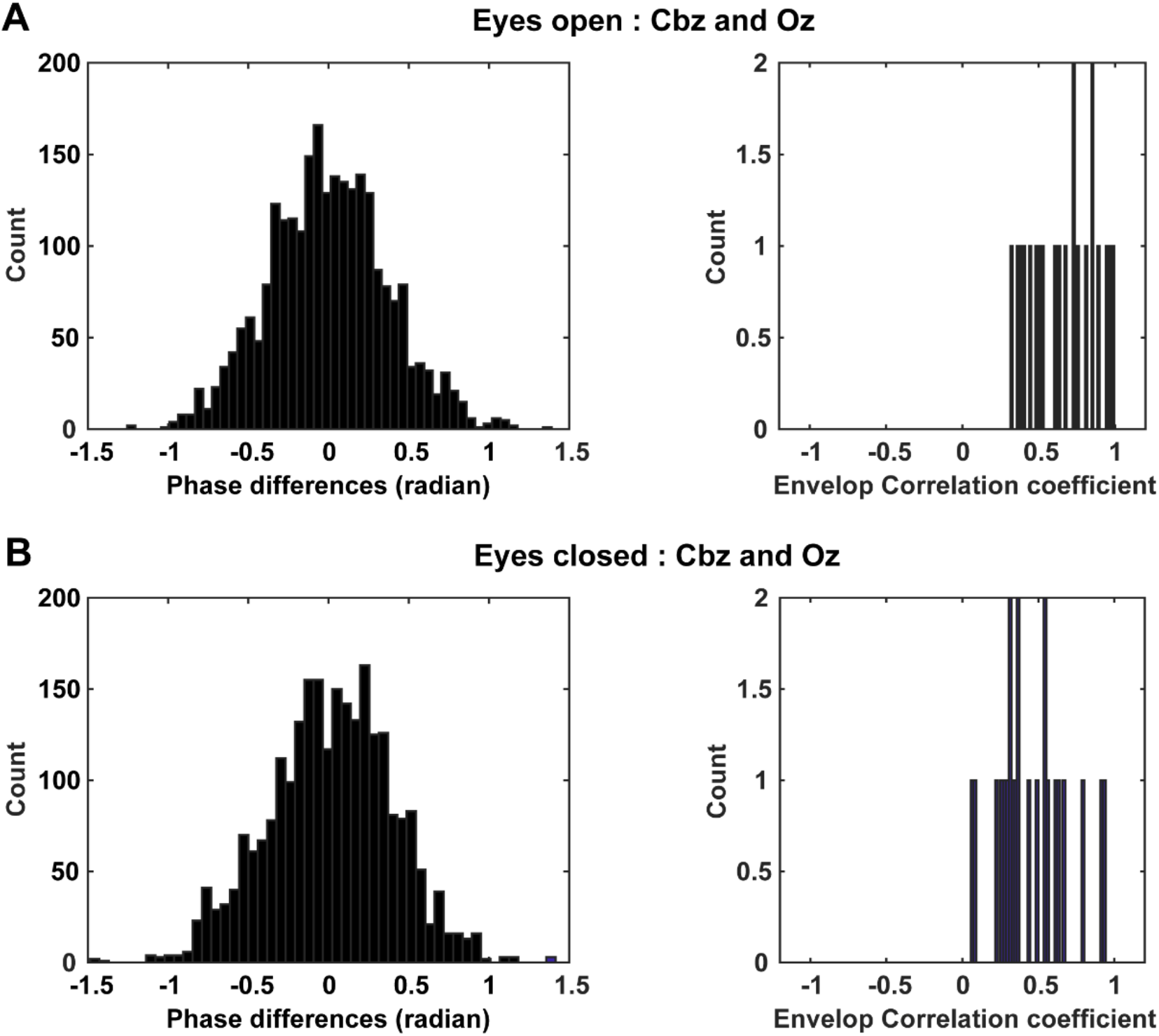
Phase and envelope analyses between Cbz and Oz (n=23 PD subjects). A-B) Average histograms of phase angle differences and average envelope correlation coefficient across 23 PD patients with 50 bins show dispersed and greater differences in phase angle, and lower envelop correlation coefficients during the resting-state eyes-open and eyes-closed conditions.

**Fig. S2.**
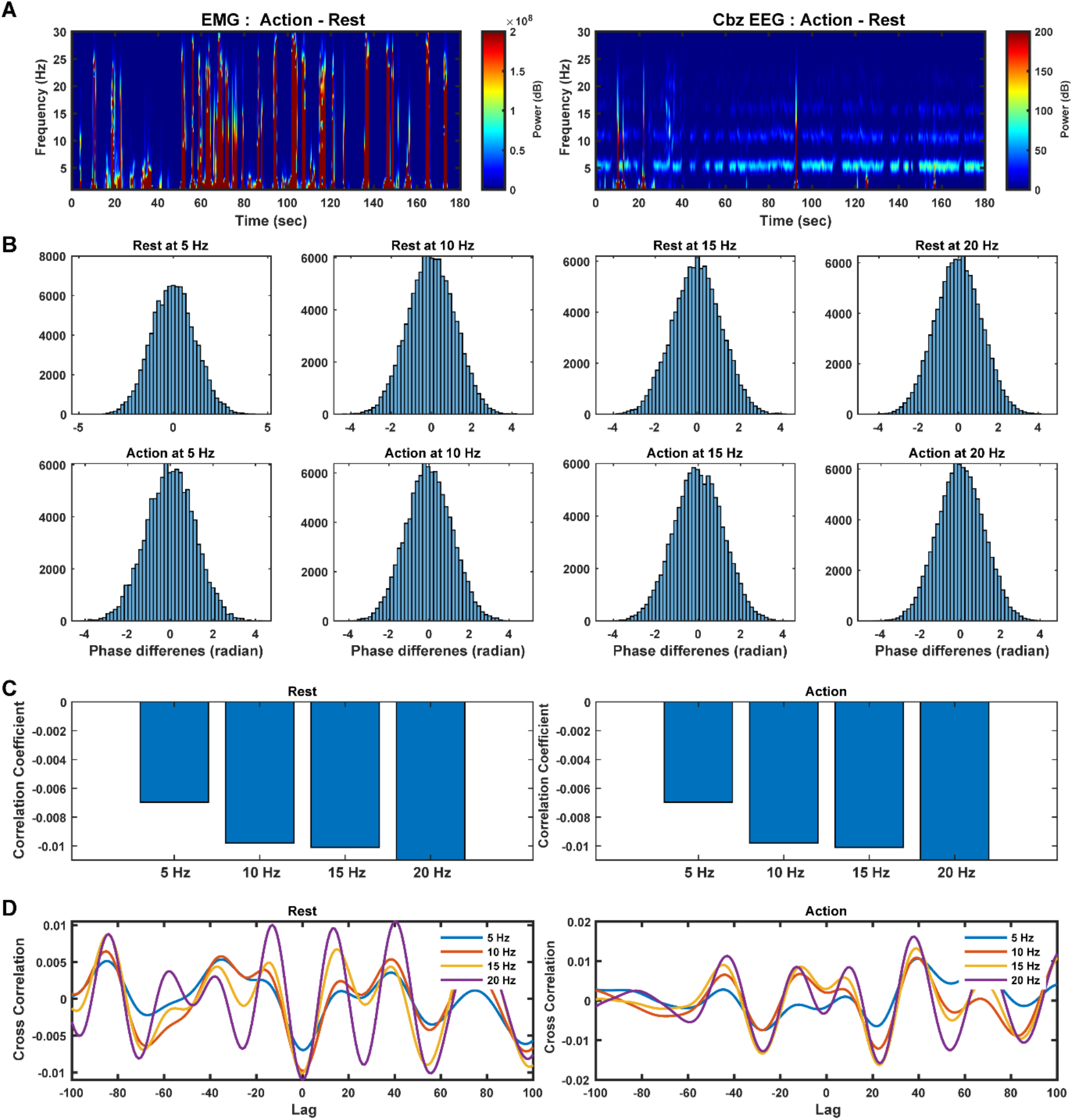
Mid-cerebellar EEG and nearby EMG signals during rest and action (n=5 PD subjects). A) Average time frequency differences between action and rest conditions for EMG electrodes placed over the semispinalis capitis muscle across 5 PD subjects. B) Average histograms with 50 bins at 5, 10, 15, and 20 Hz assessing phase differences between EMG and EEG electrodes. C) Average correlation coefficients between EEG and EMG electrodes. D) Average cross correlation analyses between EMG and EEG electrodes assessing zero-lag relationships.

**Fig. S3.**
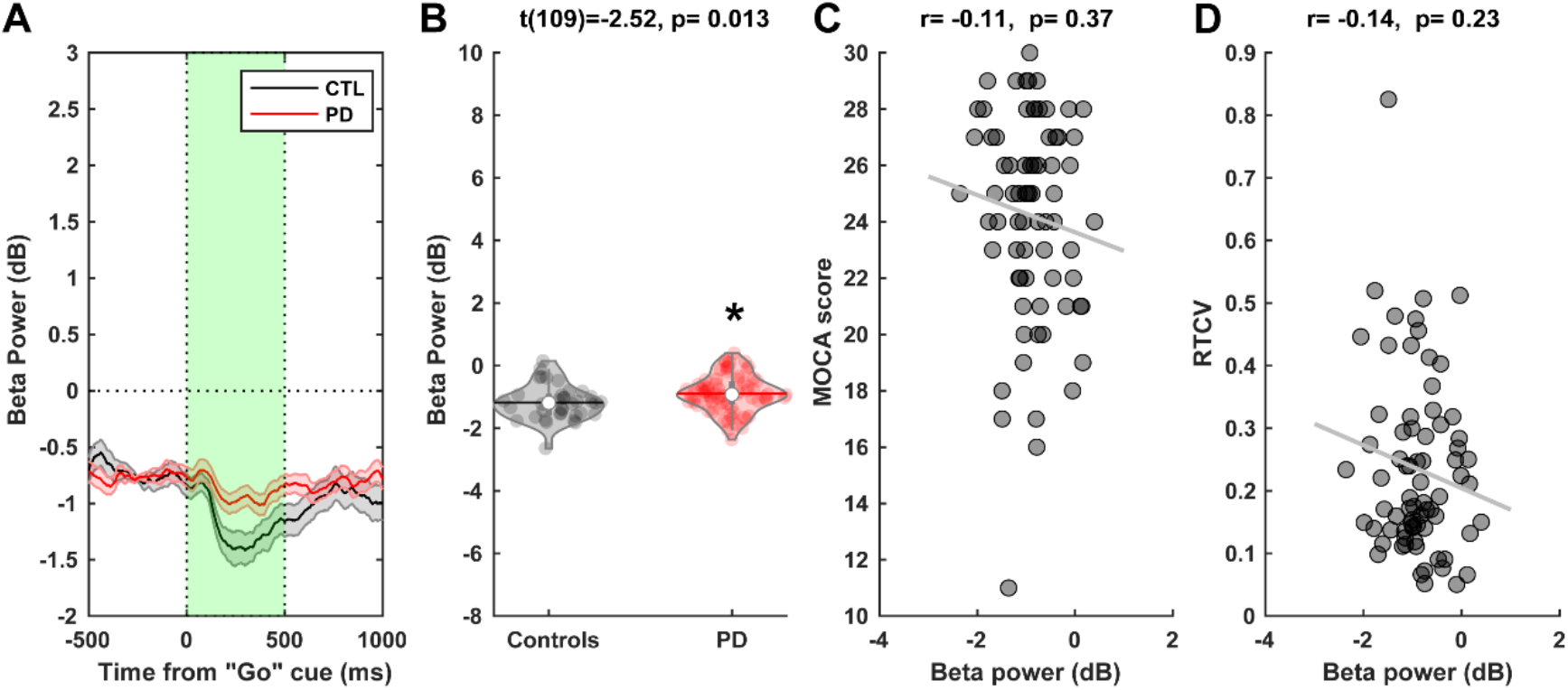
Cue-triggered mid-cerebellar Cbz beta-band activity during the interval timing task. A) Beta power (13-30 Hz) comparison between PD and control subject at the onset of the target “Go” cue. B) Beta power is significantly increased in PD patients compared to controls. C-D) No relationships is observed between beta power and cognition as indicated by MOCA scores and RTCV. *p<0.05 vs. controls. The horizontal lines and white circles in the violin plots represent the mean and median values, respectively.

**Fig. S4.**
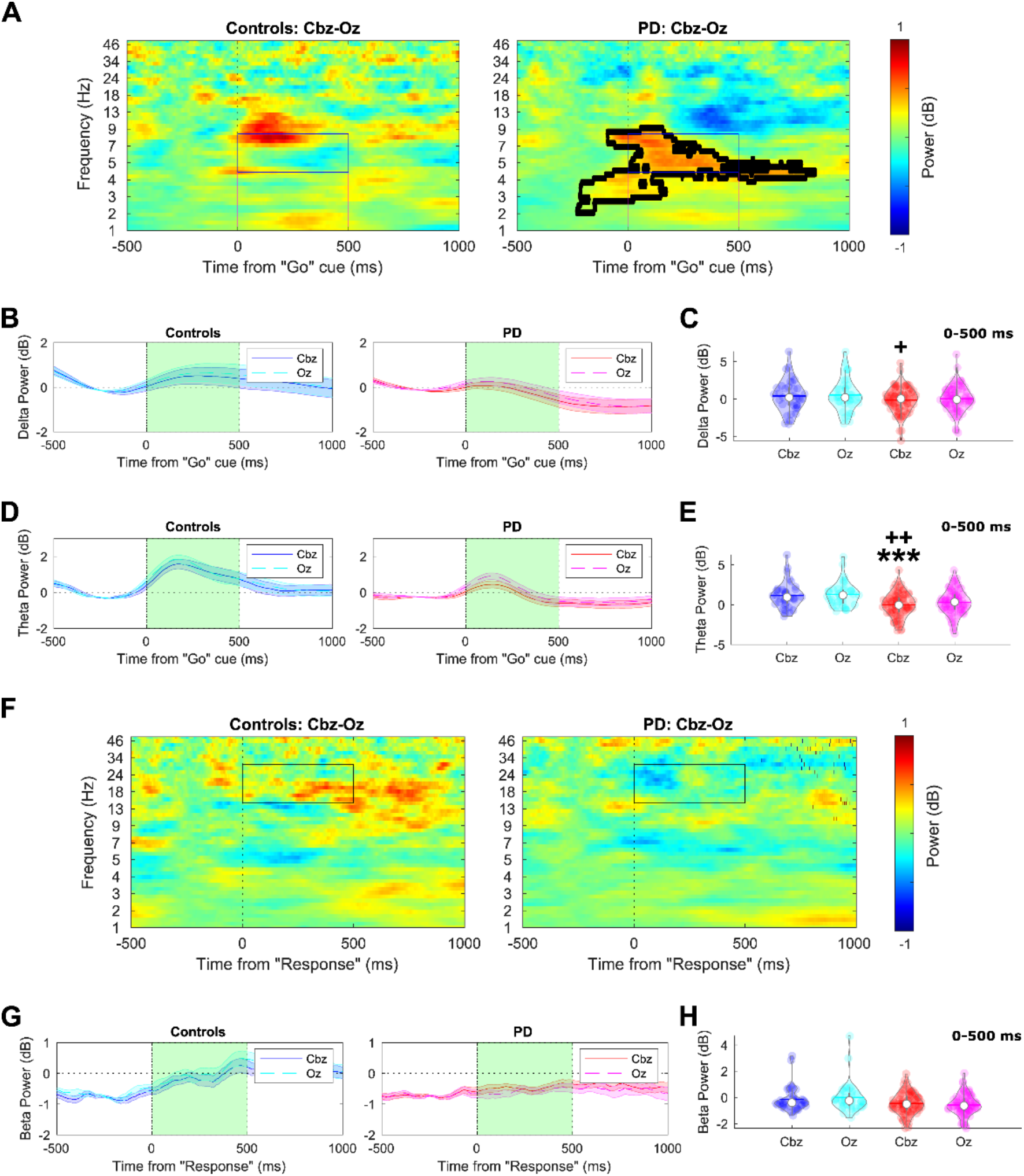
Comparison between mid-cerebellar Cbz and mid-occipital Oz delta, theta, and beta power during an interval timing cognitive task. A) Time-frequency analyses comparing cue-triggered Cbz and Oz activity in PD and controls. B-C) Cue-triggered delta is decreased in PD at Cbz. D-E) Cue-triggered theta is decreased in PD at Cbz. F) Time-frequency analyses comparing response-triggered Cbz and Oz activity in PD and controls. G-H) Response-triggered beta shows no difference. Please see table S1 for statistical analysis. +p<0.05 vs. PD-Oz; ++p<0.01 vs. PD-Oz; ***p<0.001 vs. control-Cbz. The horizontal lines and white circles in the violin plots represent the mean and median values, respectively.

**Fig. S5.**
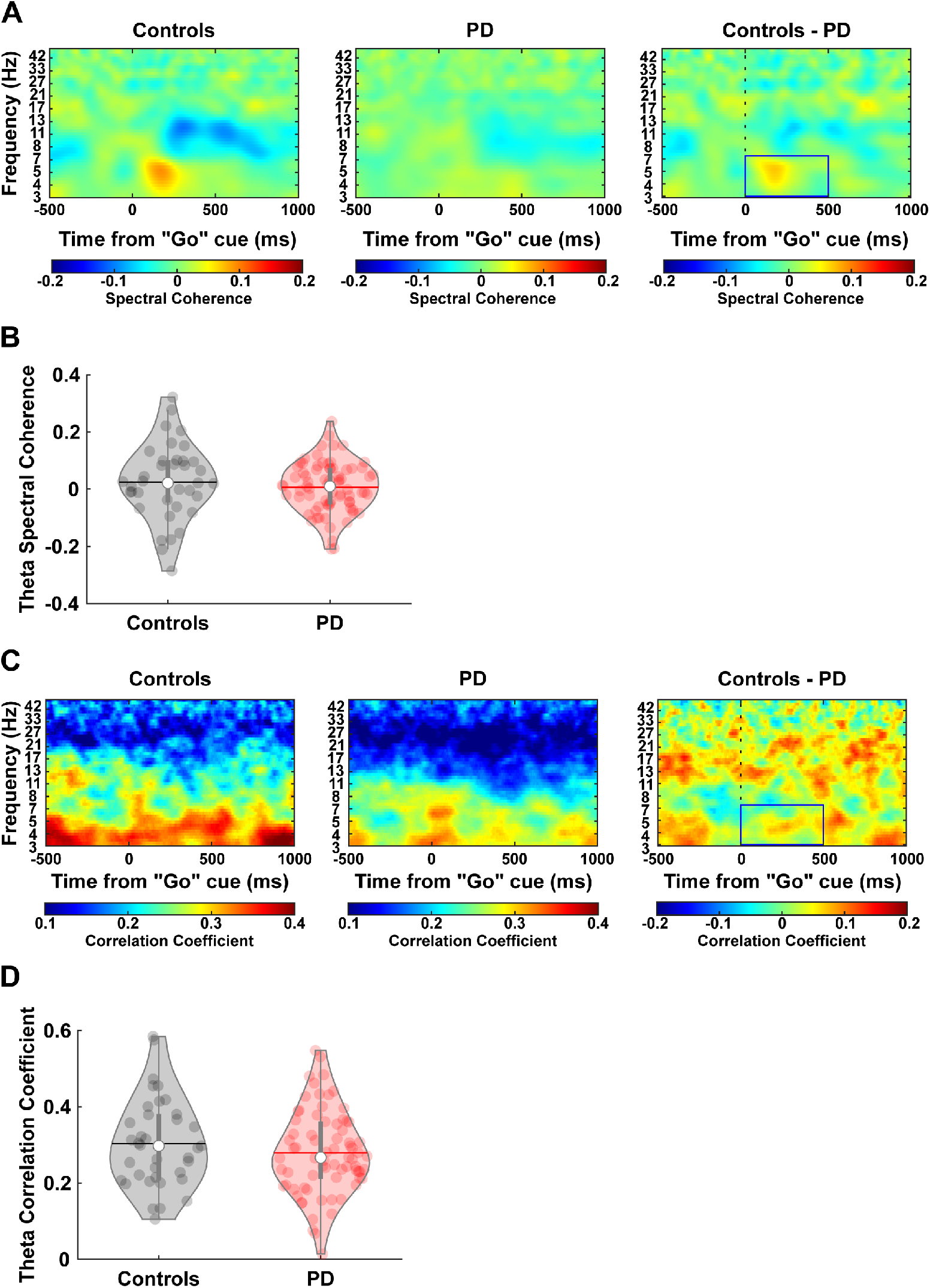
Mid-cerebellar and mid-frontal connectivity analyses comparing PD patients and control subjects during an interval timing cognitive task. (A) Time-frequency analysis comparing Cbz and Cz spectral coherence in PD and control subjects. (B) No difference in spectral coherence is observed between PD and control subjects in the theta-band. (C) Time-frequency analysis comparing Cbz and Cz power correlation coefficients in PD and control subjects. (D) No difference in power correlation coefficients were observed between PD and control subjects in the theta-band. The horizontal lines and white circles in the violin plots represent the mean and median values, respectively.

**Fig. S6.**
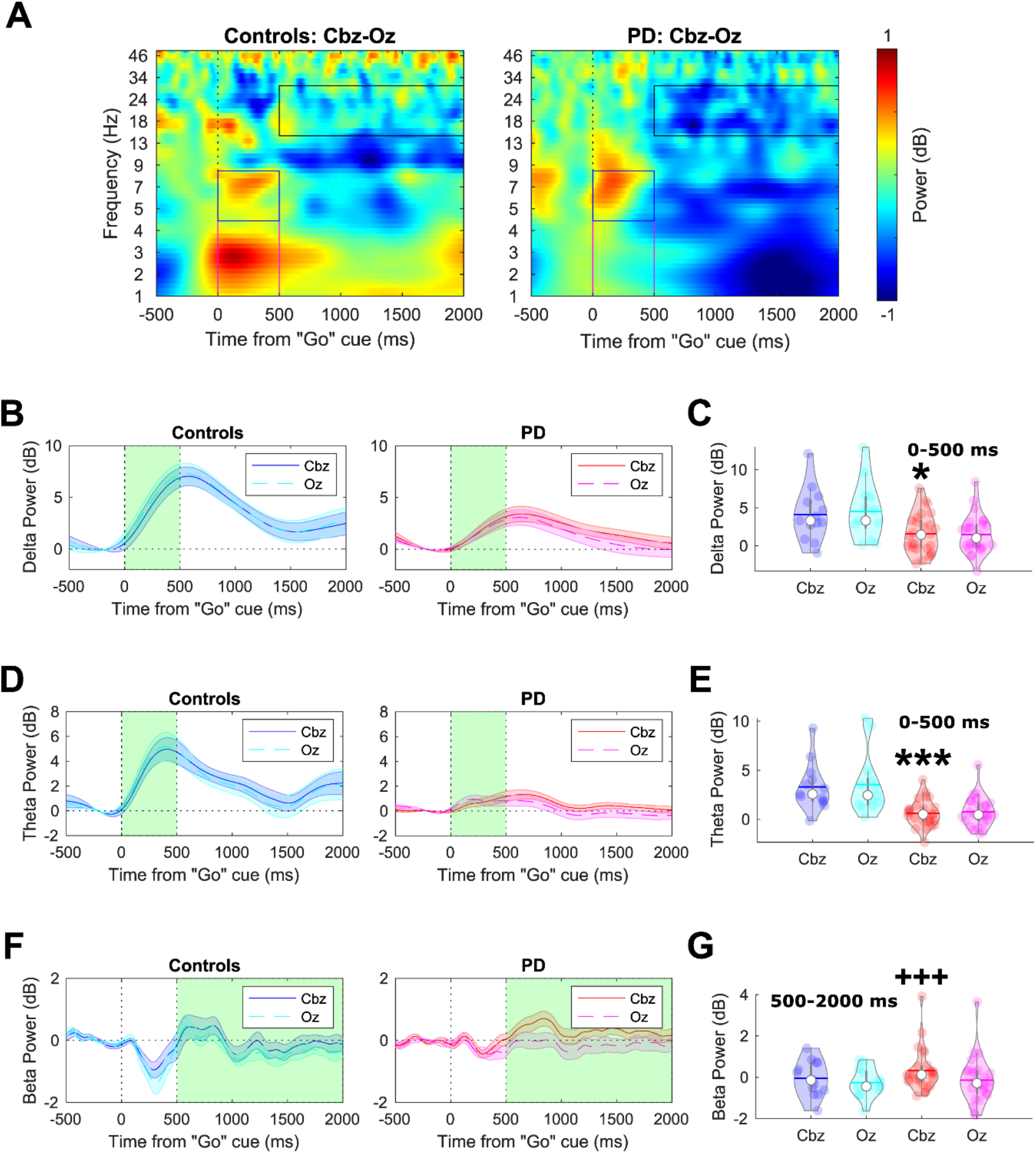
Comparison between mid-cerebellar Cbz and mid-occipital Oz delta, theta, and beta power during a lower-limb pedaling motor task. A) Time-frequency analysis comparing cue-triggered Cbz and Oz activity in PD and controls. B-C) Cue-triggered delta power is decreased in PD at Cbz. D-E) Cue-triggered theta power is decreased in PD at Cbz. F-G) Cue-triggered beta in the 500-2000 ms time-window is increased in PD at Cbz. Please see table S1 for statistical analysis. *<0.05 vs. control-Cbz; ***<0.001 vs. control-Cbz; +++p<0.001 vs. PD-Oz. The horizontal lines and white circles in the violin plots represent the mean and median values, respectively.

**Fig. S7.**
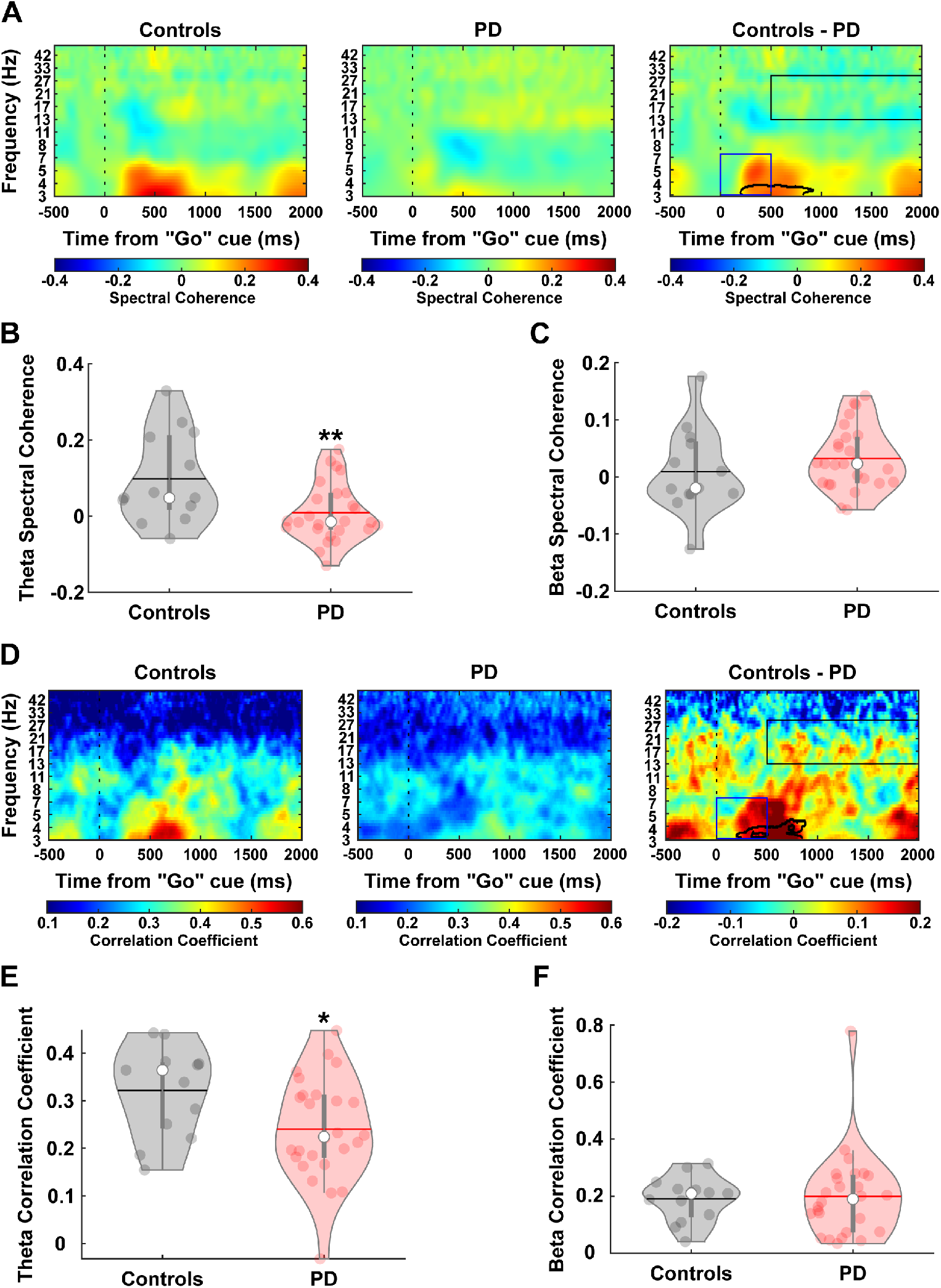
Mid-cerebellar and mid-frontal connectivity analyses comparing PD patients and control subjects during a lower-limb pedaling motor task. (A) Time-frequency analysis comparing Cbz and Cz spectral coherence in PD and control subjects. (B-C) Spectral coherence between Cz and Cbz is decreased at the theta-band in PD and no difference is observed in the beta-band. (D) Time-frequency analysis comparing Cbz and Cz power correlation coefficients in PD and control subjects. (E-F) Correlation coefficient between Cbz and Cz is decreased at the theta-band in PD and no difference is observed in the beta-band. *p<0.05 vs. controls. **p<0.01 vs. controls. The horizontal lines and white circles in the violin plots represent the mean and median values, respectively.

**Fig. S8.**
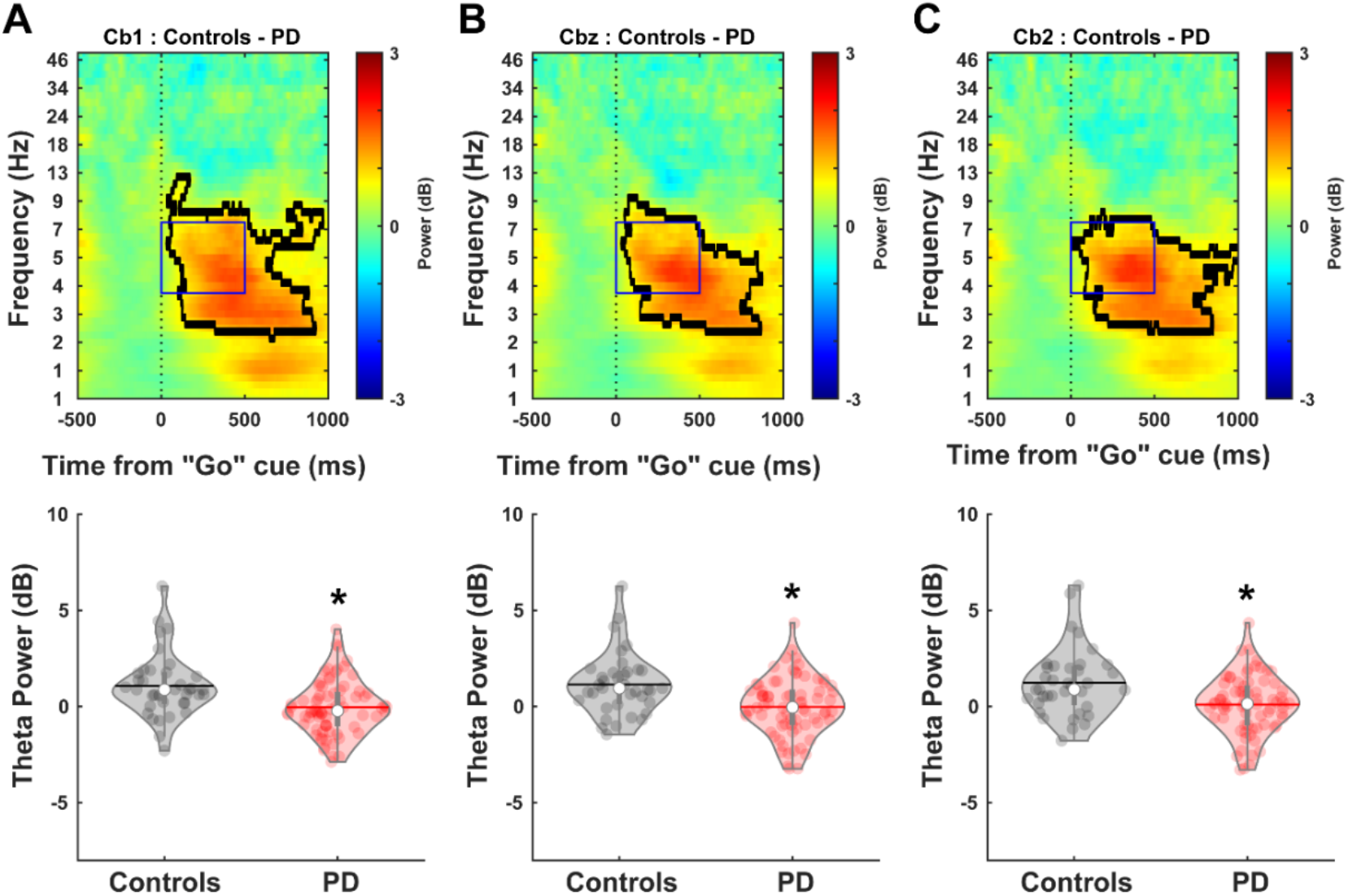
Comparison between PD and control subjects during the interval timing cognitive task at lateral cerebellar (left Cb1 and right Cb2) and mid-cerebellar Cbz locations. Time-frequency analyses show attenuated theta-band power in PD patients at A) left lateral Cb1, B) mid-cerebellar Cbz, and C) right lateral Cb2. *<0.01 vs. controls. The horizontal lines and white circles in the violin plots represent the mean and median values, respectively.

**Fig. S9.**
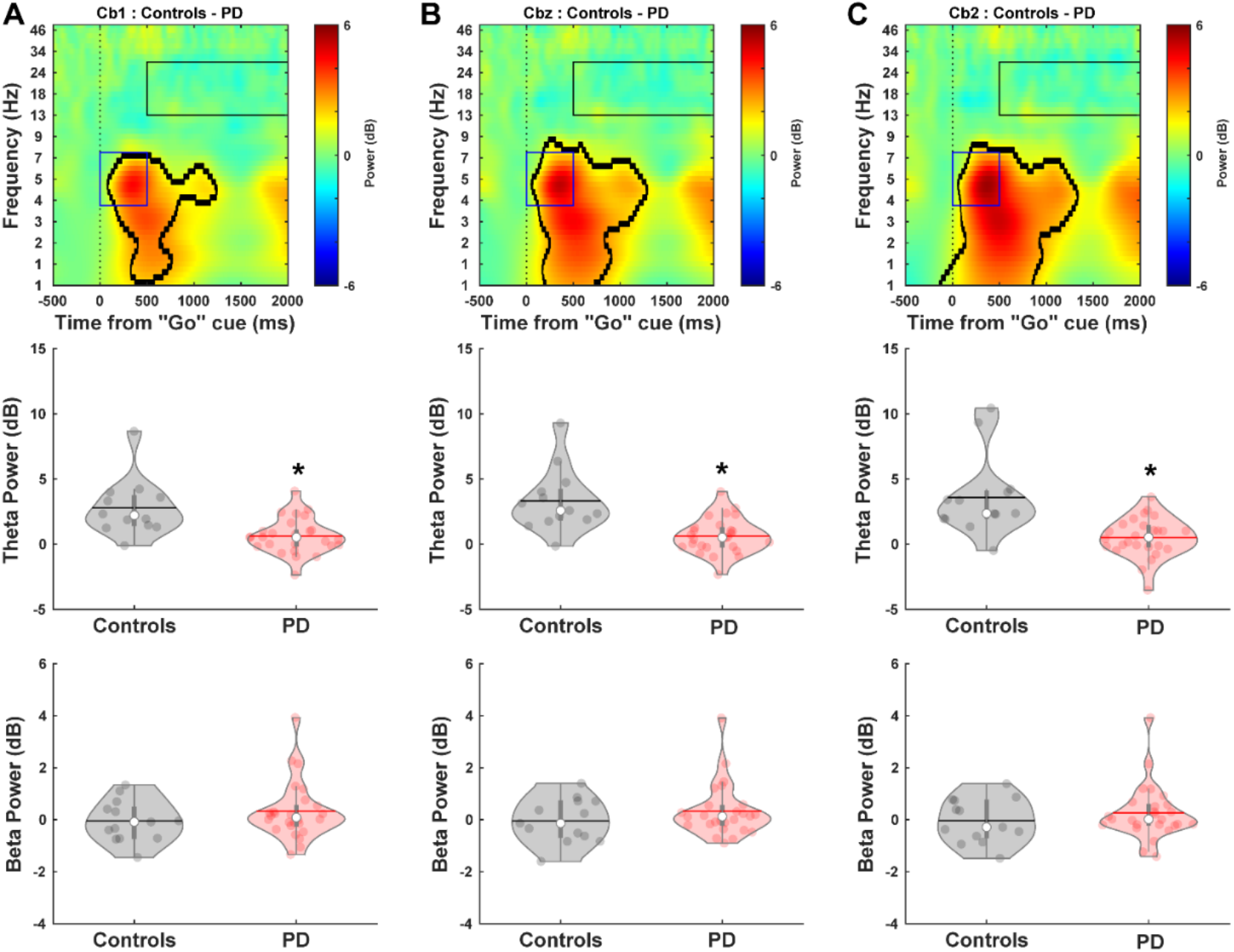
Comparison between PD and control subjects during the lower-limb pedaling motor task at lateral cerebellar (left Cb1 and right Cb2) and mid-cerebellar Cbz locations. Middle row: time-frequency analyses show attenuated theta-band power in PD patients A) left lateral Cb1, B) mid-cerebellar Cbz, and C) right lateral Cb2. Bottom row: no differences are observed in the beta-band. *<0.01 vs. controls. The horizontal lines and white circles in the violin plots represent the mean and median values, respectively.

**Table S1.** Summary of repeated measure ANOVA tests for tf-ROIs during interval timing cognitive and lower-limb pedaling motor tasks.

|  | **Group**<br>**(Controls and**<br>**PD patients)** | **Electrode**<br>**(Cbz and Oz)** | **Group\*Electrode** | **Pairwise**<br>**comparisons:**<br>**(Controls vs. PD)** | **Pairwise**<br>**comparisons:**<br>**(Cbz vs. Oz)** |
| --- | --- | --- | --- | --- | --- |
| **Interval timing cognitive task** |  |  |  |  |  |
| **Delta-band** | F(1,109)=1.97<br>p=0.16 | F(1,109)=6.83<br>p=0.01* | F(1,109)=0.15<br>p=0.7 | Cbz p=0.15<br>Oz p=0.2 | CTL p=0.15<br>PD p=0.033* |
| **Theta-band** | F(1,109)=13.2<br>p=0.0004*** | F(1,109)=8.6<br>p=0.004** | F(1,109)=0.89<br>p=0.35 | Cbz p<0.0001***<br>Oz p=0.002** | CTL p=0.39<br>PD p=0.003** |
| **Beta-band** | F(1,109)=6.65<br>p=0.01* | F(1,109)=0.35<br>p=0.55 | F(1,109)=6.7<br>p=0.01* | Cbz p=0.07<br>Oz p=0.003** | CTL p=0.068<br>PD p=0.058 |
| **Lower-limb pedaling motor task** |  |  |  |  |  |
| **Delta-band** | F(1,37)=8.1<br>p=0.007** | F(1,37)=0.17<br>p=0.68 | F(1,37)=1.35<br>p=0.25 | Cbz p=0.016*<br>Oz p=0.005* | CTL p=0.045*<br>PD p=0.77 |
| **Theta-band** | F(1,37)=17.4<br>p=0.0002*** | F(1,37)=1.5<br>p=0.22 | F(1,37)=0.11<br>p=0.74 | Cbz p<0.0001***<br>Oz p=0.001** | CTL p=0.48<br>PD p=0.3 |
| **Beta-band** | F(1,37)=0.59<br>p=0.45 | F(1,37)=17.1<br>p=0.0002*** | F(1,37)=1.64<br>p=0.21 | Cbz p=0.27,<br>Oz p=0.72 | CTL p=0.22<br>PD p<0.0001*** |

\*\*\*p<0.001, \*\*p<0.01, \*p<0.05.

**Table S2.** Mean ± standard error of mean for cerebellar electrodes.

|  | **Cb1 electrode** | **Cbz electrode** | **Cb2 electrode** |
| --- | --- | --- | --- |
| *Interval Timing Cognitive Task* |  |  |  |
| *Theta Band* |  |  |  |
| PD | -0.05 ± 0.16 | -0.01 ± 0.17 | 0.1 ± 0.17 |
| Controls | 1.08 ± 0.27 | 1.15 ± 0.26 | 1.24 ± 0.29 |
| *Lower-Limb Pedaling Motor Task* |  |  |  |
| *Theta Band* |  |  |  |
| PD | 0.63 ± 0.27 | 0.62 ± 0.27 | 0.5 ± 0.3 |
| Controls | 2.77 ± 0.6 | 3.31 ± 0.67 | 3.58 ± 0.85 |
| *Beta Band* |  |  |  |
| PD | 0.32 ± 0.22 | 0.32 ± 0.2 | 0.26 ± 0.21 |
| Controls | -0.05 ± 0.22 | -0.04 ± 0.24 | -0.04 ± 0.24 |

**Table S3.**
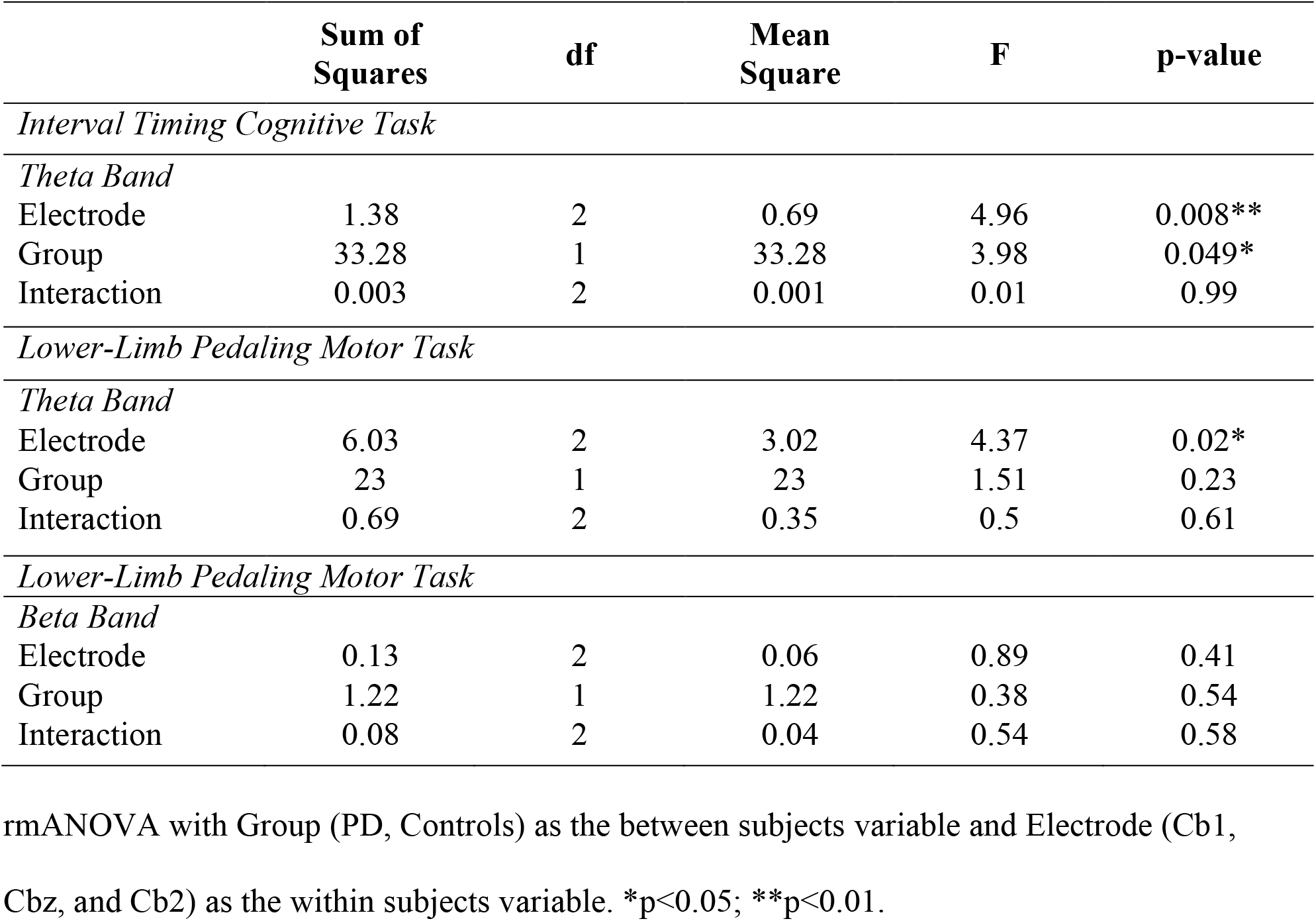
Summary of repeated measure ANOVA tests (Cb1, Cbz, and Cb2 electrodes/controls, PD groups) for tf-ROIs during interval timing cognitive and lower-limb pedaling motor tasks.

|  | **Sum of Squares** | **df** | **Mean Square** | **F** | **p-value** |
| --- | --- | --- | --- | --- | --- |
| *Interval Timing Cognitive Task* |  |  |  |  |  |
| *Theta Band* |  |  |  |  |  |
| Electrode | 1.38 | 2 | 0.69 | 4.96 | 0.008** |
| Group | 33.28 | 1 | 33.28 | 3.98 | 0.049* |
| Interaction | 0.003 | 2 | 0.001 | 0.01 | 0.99 |
| *Lower-Limb Pedaling Motor Task* |  |  |  |  |  |
| *Theta Band* |  |  |  |  |  |
| Electrode | 6.03 | 2 | 3.02 | 4.37 | 0.02* |
| Group | 23 | 1 | 23 | 1.51 | 0.23 |
| Interaction | 0.69 | 2 | 0.35 | 0.5 | 0.61 |
| *Lower-Limb Pedaling Motor Task* |  |  |  |  |  |
| *Beta Band* |  |  |  |  |  |
| Electrode | 0.13 | 2 | 0.06 | 0.89 | 0.41 |
| Group | 1.22 | 1 | 1.22 | 0.38 | 0.54 |
| Interaction | 0.08 | 2 | 0.04 | 0.54 | 0.58 |

rmANOVA with Group (PD, Controls) as the between subjects variable and Electrode (Cb1, Cbz, and Cb2) as the within subjects variable. \* $p < 0.05$ ; \*\* $p < 0.01$ .

